# Job Insecurity and Proxy Suicide Risk in UK Adults: A Longitudinal Analysis of Social Determinants

**DOI:** 10.1101/2025.08.15.25333771

**Authors:** Eleftheria Vaportzis

**Affiliations:** Department of Psychology, University of Bradford, Bradford, UK

**Author notes:** Corresponding author details: Full name Eleftheria Vaportzis, Postal address Department of Psychology, University of Bradford, Bradford, BD7 1DP, UK.

**Keywords:** job insecurity, suicide risk, social determinants, longitudinal analysis, subjective wellbeing, loneliness

## Abstract

**Introduction:** Job insecurity is a known risk factor for suicide. The extent to which perceived job insecurity impacts suicide risk is unclear. This study examined whether perceived job insecurity is associated with suicide risk, operationalised via a proxy score derived from variables empirically linked to suicide risk, and whether this association is confounded by broader social determinants.

**Method:** Data were drawn from Waves 10, 12, and 14 of the UK Household Longitudinal Study, comprising 33,364 observations from 20,566 participants between 18 and 66 years old. A proxy for suicide risk was constructed using the following five variables: subjective wellbeing (GHQ-12), loneliness, general health, life satisfaction, and subjective financial situation. Job insecurity was measured via self-reported likelihood of job loss. Generalised Estimating Equations were used to model associations, adjusting for sex, ethnicity, marital status, education, and income.

**Results:** Unadjusted models showed a significant association between perceived job insecurity and suicide risk. However, in a multivariable model controlling for social determinants including low income, being divorced or separated, and lower education, this association was no longer significant. On the contrary, being male, married or cohabiting, or holding a university degree were associated with lower odds of suicide risk.

**Conclusion:** Although perceived job insecurity initially appeared to be associated with suicide risk, its effect is largely explained by social determinants. These findings highlight the importance of integrating social determinants into suicide prevention strategies. Targeting financial hardship, relationship instability, and educational disadvantage may be more effective than addressing job insecurity alone. The use of a validated proxy score offers a feasible and scalable approach for assessing suicide risk in large-scale datasets lacking direct measures.

---

Suicide is a public health concern that arises from social, cultural, biological, psychological, and environmental factors present across the life-course, and results in over 720,000 deaths every year (WHO, 2025). The long-term association between suicide risk and social determinants of health, including income, social protection, and unemployment, is well-established (Gallagher et al., 2025). Beyond the crude distinction between employment versus unemployment, subjective perceptions of job insecurity (i.e., the fear of losing one’s job) have a negative impact on people’s wellbeing and work engagement (Pires, 2025). Job insecurity may pose a threat comparable to that of unemployment, contributing to mental and general health symptoms such as depression, anxiety, and cardiovascular disease (Kim & von Dem Knesebeck, 2015).

Job insecurity has been linked to an increased risk of suicidal outcomes, including suicidal ideation, self-harm, and suicide attempts (Mathieu, Treloar, Hawgood, Ross, & Kõlves, 2022; Waterall, Newland, & Murphy, 2022). Longitudinal studies have shown that declines in job security are associated with declines in mental health; these declines are moderated by social determinants such as income, marital status and education suggesting that the psychological impact of job insecurity is affected by the broader socioeconomic context (Gallagher et al., 2025; Thomson, Kopasker, Leyland, Pearce, & Katikireddi, 2022).

Durkheim’s classic work on suicide (Durkheim, 2005) posits that suicide rates are related to how communities provide their members with a sense of purpose that makes life worth living even in stressful times. Employment plays a crucial role in how individuals experience their communities, derive a sense of purpose in life, and consequently develop a sense of wellbeing (Sandoghdar & Grimshaw, 2023). In relation to job insecurity, Durkheim’s theory considers social determinants (e.g., income, marital status, education) not only to confound the association between stress related to job insecurity and suicide risk but also to influence individuals’ experience of stress and how they cope with it. While Durkheim emphasises macro-level social integration, Abrutyn and Mueller (2014) have highlighted how socioemotional disruptions at the individual level (e.g., relationship difficulties) may mediate the effects of broader social determinants.

Research exploring the association between job insecurity and suicide provides mixed evidence. A meta-analysis of studies showed that job-related stress was related to suicidal outcomes (Milner, Witt, LaMontagne, & Niedhammer, 2018). Blomqvist, Virtanen, LaMontagne, and Magnusson Hanson (2022) used cohort data from a Swedish occupational survey linked to national registers and reported that perceived job insecurity predicted suicide deaths. Follow-up analyses during the COVID-19 pandemic suggested a stronger association for actual job loss than perceived job insecurity and suicidal ideation (Blomqvist, Westerlund, & Hanson, 2024). Controlling for personality traits reduced the effect of job insecurity, suggesting that other factors may confound this association. Similarly, Mathieu et al. (2022) reported that while job insecurity was associated with suicidal behaviours, the strength of association decreased once income, education, and marital status were accounted for.

Despite the growing literature on the topic of job insecurity and suicide, important gaps remain. Although cohort studies suggest that job insecurity is associated with increased risk of suicidal outcomes, it remains unclear whether perceived job insecurity independently predicts suicide risk once social determinants are accounted for. Recent studies have reported a weakening of this relationship after adjusting for factors such as income, marital status, and education (Blomqvist, Virtanen, LaMontagne, & Magnusson Hanson, 2022; Blomqvist, Westerlund, & Hanson, 2024). In addition, although many longitudinal studies lack validated measures of suicidal behaviours, proxy scores derived from variables empirically linked to suicide risk can offer a robust alternative when carefully constructed and validated.

The present study uses longitudinal data from the UK Household Longitudinal Study (UKHLS) to examine whether perceived job insecurity is associated with suicide-risk operationalised through a proxy score drawn from variables that have been linked to higher suicide risk (e.g., subjective wellbeing, loneliness). The study also examines whether this relationship is moderated by social determinants (e.g., income, social status). The aim is to understand the extent to which perceived job insecurity is associated with suicide risk and whether this association is accounted for by broader social determinants.

## Methods

### Primary data source

We obtained the data from the UKHLS (www.understandingsociety.ac.uk; University of Essex, 2024). In this paper, we summarise key aspects of the survey’s development and its methodology. A comprehensive account can be found in several other reports (e.g., Buck & McFall, 2011; McFall & Garrington, 2011).

UKHLS is a longitudinal study interviewing everyone in a household to see how different generations experience life in the UK. Wave 1 of data collection was completed between January 2009 and December 2011. Sampling was done from the Postcode Address File in Great Britain and the Land and Property Services Agency list of domestic properties in Northern Ireland, and identified 55,684 eligible households. Interviews were completed with a total of 50,994 participants aged 16 or older from 30,117 households. The current study uses data from Waves 10, 12, and 14. Wave 14 includes a general population sample boost (Connett, 2024).

### Variables

The outcome variable was a proxy for suicide risk. This approach is in line with previous studies that used suicide proxies in the absence of specific variables regarding suicide risk (Borges et al., 2006; Carmel, Ries, West, Bumgardner, & Roy-Byrne, 2016). UKHLS does not collect data specific to suicide risk; however, it collects data on various variables that have been linked to higher suicide risk. The suicide risk proxy was created using the following five variables:

1. Subjective wellbeing was measured with the General Health Questionnaire (GHQ-12; Goldberg & Williams, 1988). GHQ-12 comprises 12 items concerning symptoms such as general happiness, confidence, overcoming difficulties, and enjoying day-to-day activities over the past four weeks. Scores range from 0 to 12, with higher scores indicating greater psychological distress and lower subjective wellbeing;
2. Loneliness was measured with the question “How often do you feel lonely?” Responses were recorded on a 3-point Likert scale (1 = “Never” – 3 = “Often”);
3. General health was self-reported. Participants were asked, “In general, would you say your health is…” Responses were recorded on a 5-point Likert scale (1 = “Excellent” – 5 = “Poor”);
4. Life satisfaction was measured with the question “How satisfied are you currently with your life overall?”. Responses were recorded on a 7-point Likert scale (1 = “Completely dissatisfied” – 7 “Completely satisfied”). We reverse-scored this measure to ensure that higher scores consistently reflected higher risk across all measures (i.e., 1 = “Completely satisfied” – 7 = “Completely dissatisfied”); and
5. The current subjective financial situation of participants was measured with the question “How well would you say you are managing financially these days?” Responses were recorded on a 5-point Likert scale (1 = “Living comfortably” – 5 = “Finding it very difficult”).

An exploratory factor analysis using Principal Axis Factoring provided support for averaging the five variables into a proxy score for suicide risk. Kaiser-Meyer-Olkin Measure of Sampling Adequacy was .729, and Bartlett’s test of sphericity was significant (*p* < .001), suggesting that the correlation matrix was suitable for factor analysis. One factor was extracted, accounting for 30.6% of the variance. All five variables loaded positively onto the extracted factor, with loadings ranging from .44 (financial situation) to .75 (life satisfaction), suggesting they reflect a common underlying construct and supporting the unidimensionality of the proxy score.

The five variables were standardised using Z-scores, and their average created a total proxy for suicide risk. The overall Cronbach’s alpha was acceptable (α = 0.72). To assess internal consistency across waves, Cronbach’s alpha was also calculated for each wave separately (α ranged between 0.71 and 0.73). Although the individual Z-scores were almost normally distributed, the proxy score was moderately positively skewed (0.74), suggesting concentration of lower scores. The low kurtosis score (0.43) suggests a relatively flat distribution, with fewer extreme values than a normal curve. These values support the use of our dataset’s percentile cut-offs to classify the proxy scores into groups. Participants falling within the lowest 20% of scores were classified as “low risk,” those within the highest 20% as “high risk,” and the remaining middle 60% as “medium risk.”

The predictor variable was job insecurity, measured with the question “I would like you to think about your employment prospects over the next 12 months. Thinking about losing your job by being sacked, laid off, made redundant, or not having your contract renewed. How likely do you think it is that you will lose your job during the next 12 months?” Responses were recorded on a 4-point Likert scale (1 = “Very likely” – 4 = “Very unlikely”). We reverse-scored this measure to ensure that higher scores consistently reflected higher risk across all measures (i.e., 1 = “Very unlikely” – 4 = “Very likely”). We note that initially redundancy status was considered as a separate predictor variable; however, due to its extremely low frequency (< 1%), it was not included in the analysis.

Based on past research (Berkelmans, van der Mei, Bhulai, & Gilissen, 2021), we considered the following covariates: age (continuous), sex (female, male), ethnicity (e.g., White British, Indian, African), marital status (e.g., married, divorced, widowed), highest obtained educational qualification (e.g., degree, A level, GCSE), and total net personal income (quartiles).

### Sample selection

The analytic sample comprised participants aged between 18 and 66 who participated in at least one wave from Waves 10, 12, and 14 (see Table 1). Demographic information is presented for participants’ first wave of participation across all waves; therefore, the baseline sample comprises all unique participants who participated in at least one wave. Descriptive statistics are given for all participants and proxy suicide risk subgroups: “high”, “medium”, and “low risk”. One-way ANOVAs were used to compare the variables between the three subgroups.

**Table 1.**
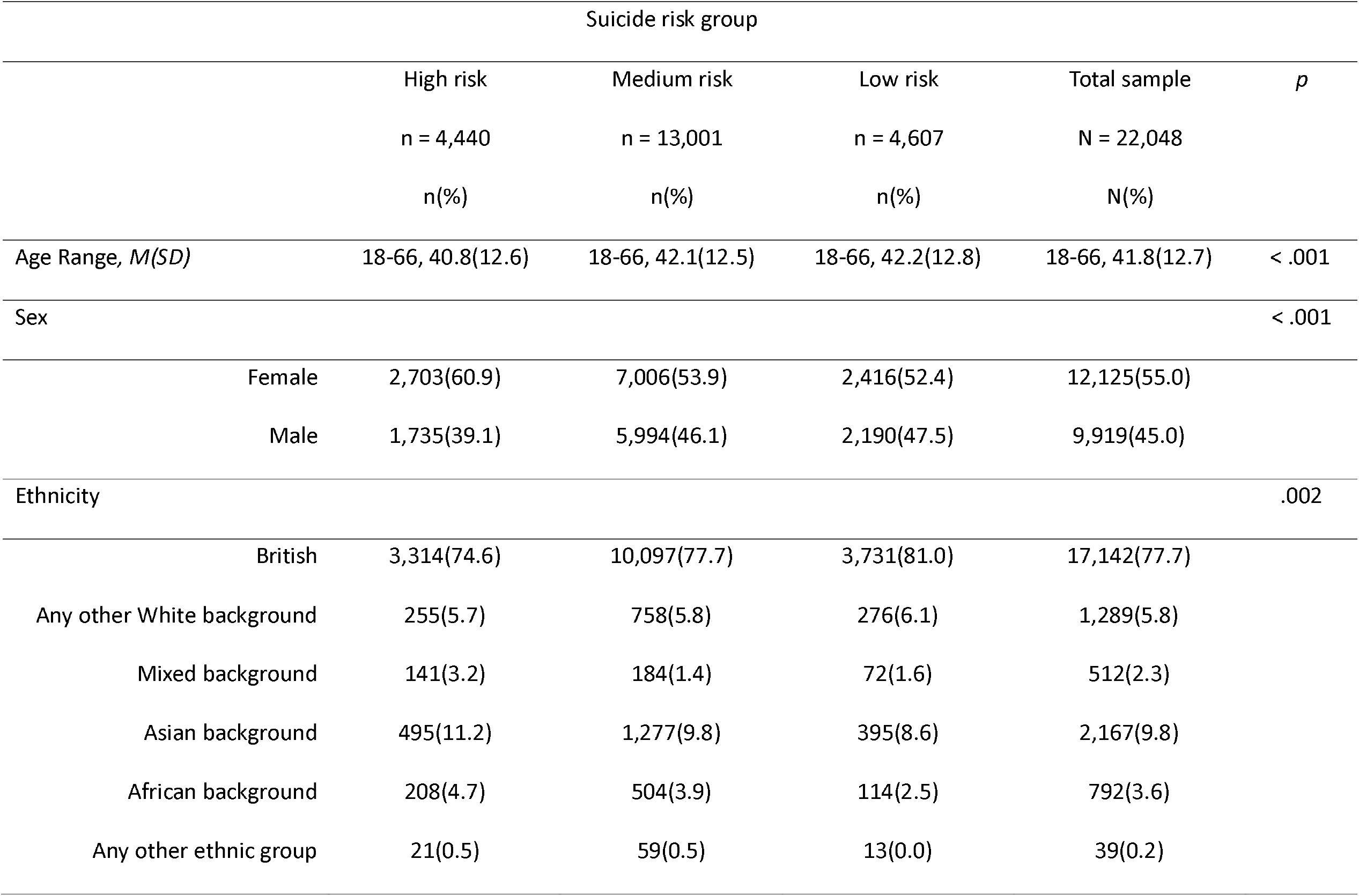
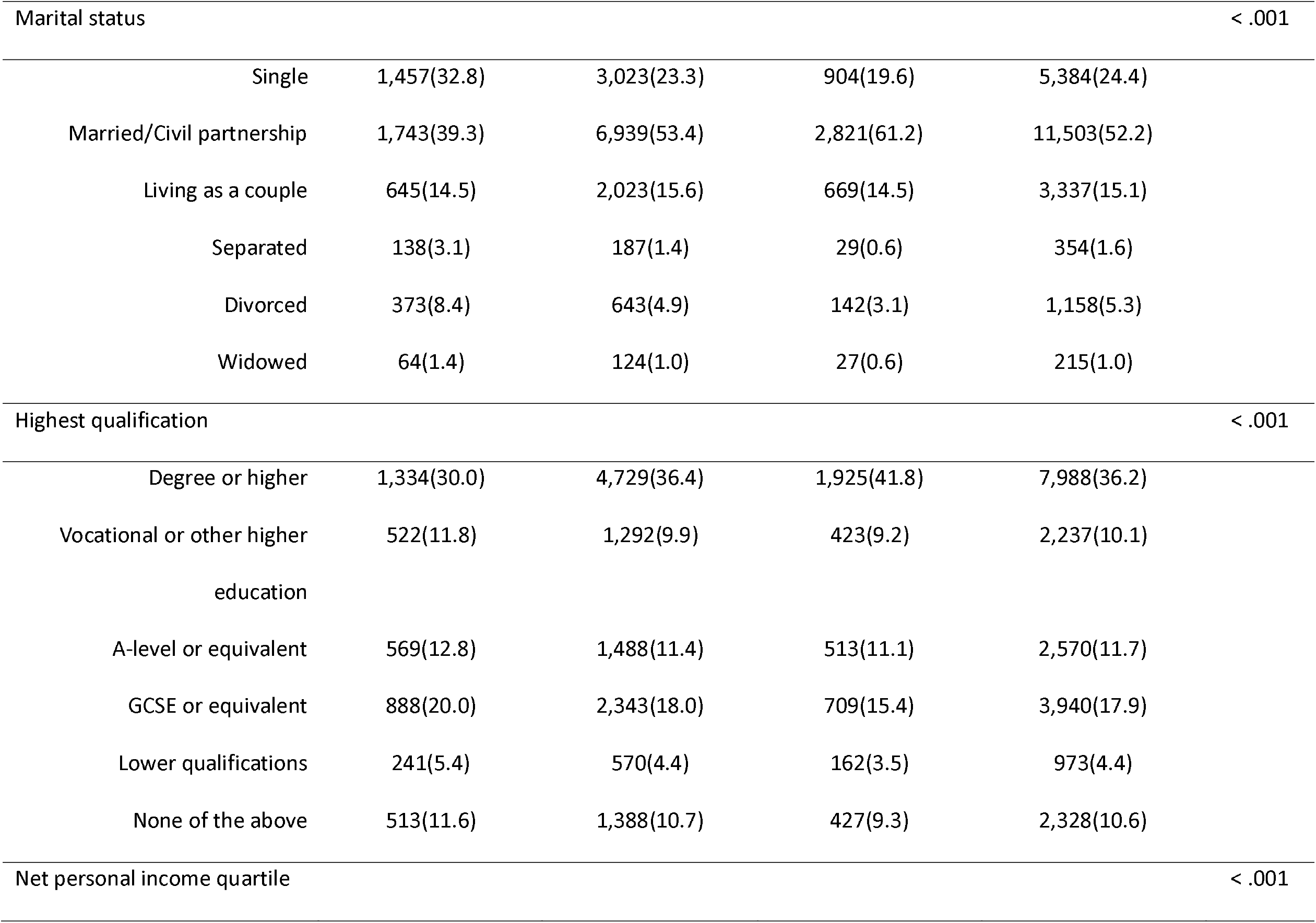
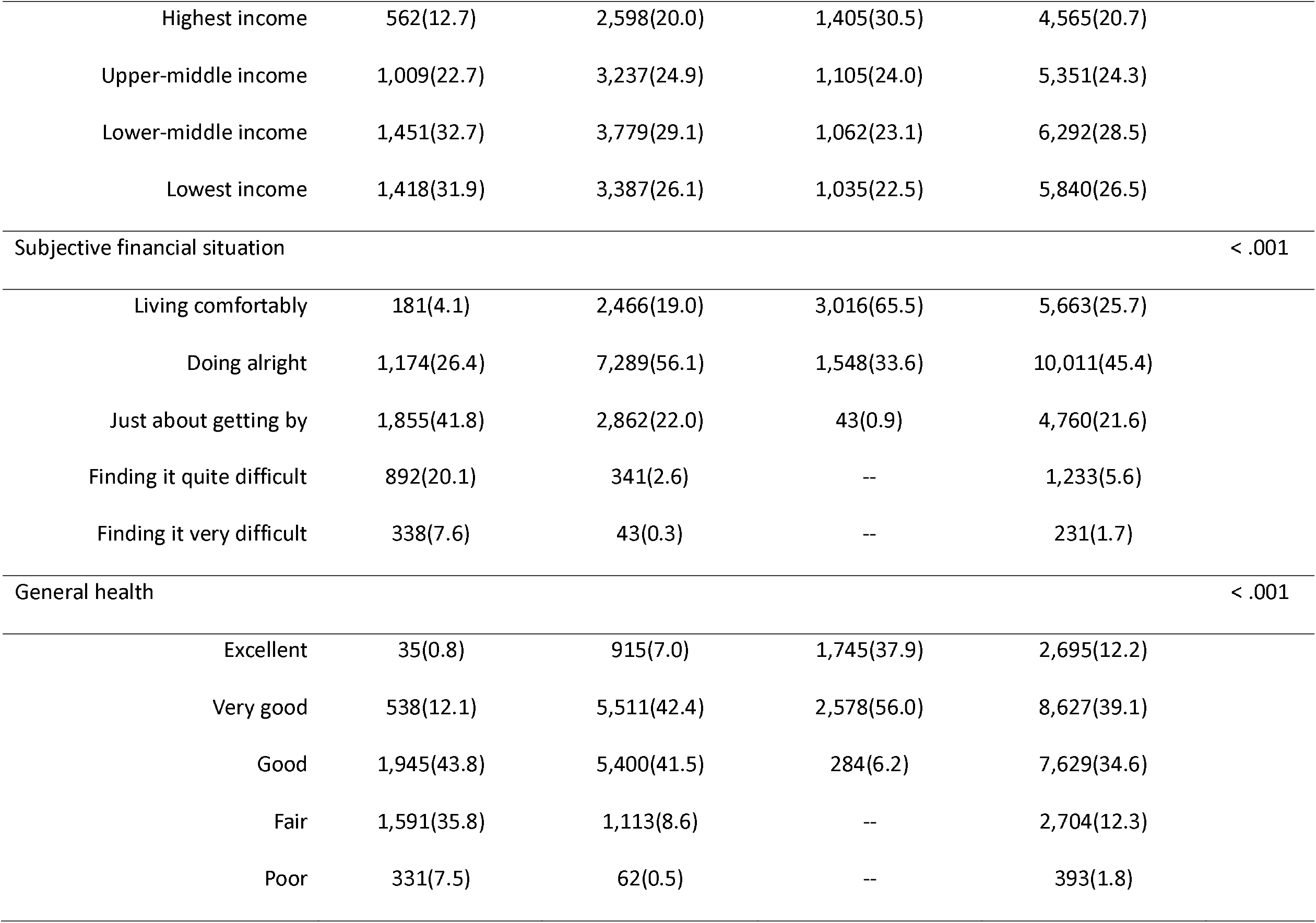
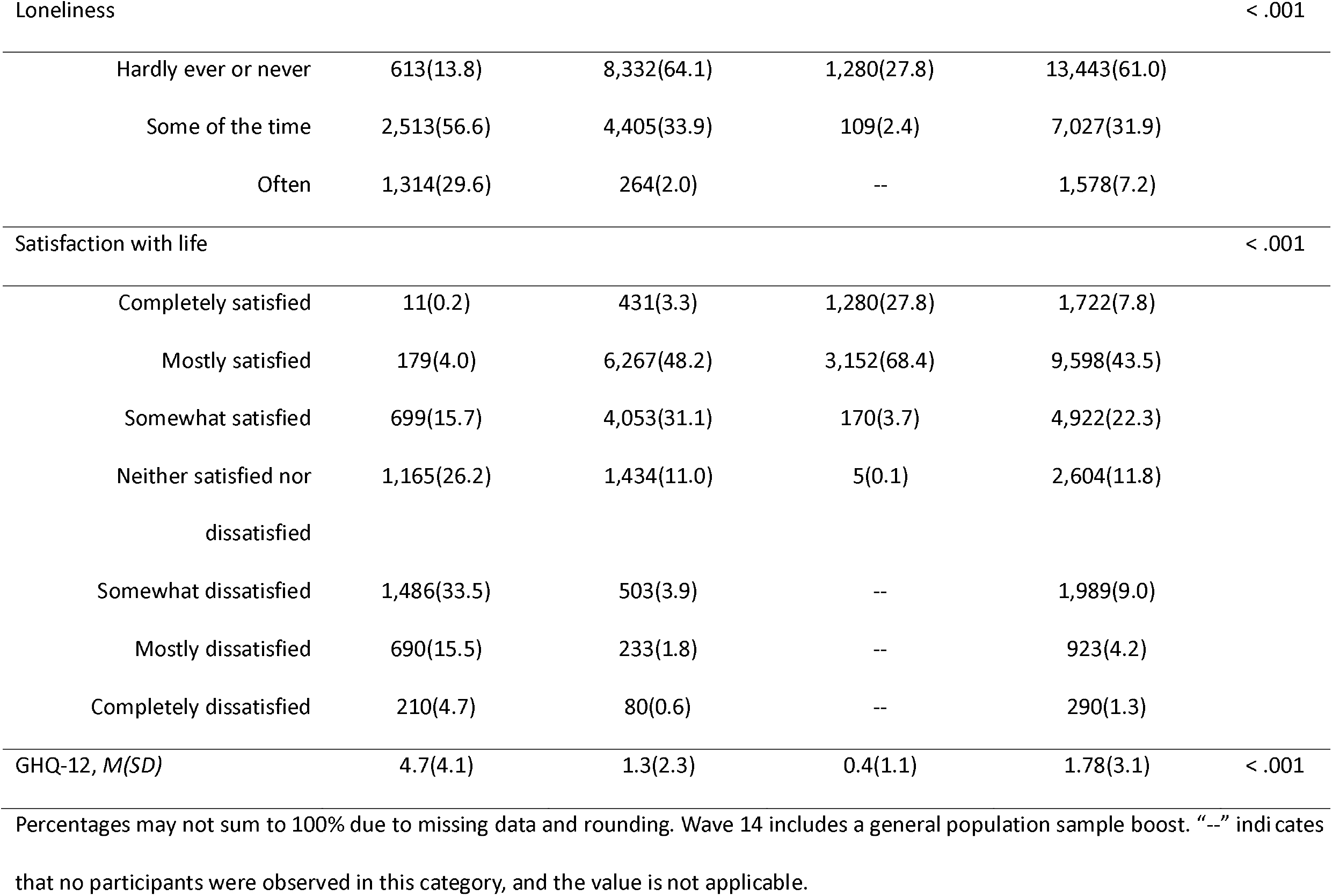
Descriptive information by suicide risk group at baseline.

### Missing data

The original sample across the three waves included 52,324 observations from 27,256 participants between 18-66 years old. We excluded listwise participants who missed all data across a wave (50,833 observations, N= 26,420) and those who missed at least one of the five core proxy variables (44,930 observations, N = 25,809). Participants who were excluded did not provide sufficient data to allow imputation; therefore, listwise deletion was deemed the most appropriate (Jakobsen, Gluud, Wetterslev, & Winkel, 2017). For job insecurity, 11,566 observations (25.7%) were missing. Due to the large number of cases missing job insecurity data, logistic binary regression analyses using age, sex, ethnicity, marital status, education, and net personal income were run to test for cases missing at random. Some of the variables were non-significant, suggesting that job insecurity data were missing at random (Little & Rubin, 2019). A sensitivity analysis compared models with and without cases missing job insecurity data. As the results were comparable, we conducted an analysis using complete-case data (Jakobsen et al., 2017). The total sample comprised 33,364 observations from 20,566 participants (14,166 in Wave 10, 882 in Wave 12, and 5,518 in Wave 14).

### Data analysis

All analyses were conducted using SPSS v.28 (IBM, 2021). Alpha was set at .05. Multicollinearity was assessed using variance inflation factors (VIFs). There was no significant collinearity among the predictors (all VIFs < 5.0). Generalised Estimating Equation (GEE) models with multinomial distribution and an ordinal logistic link were employed to examine the association between job insecurity and suicide risk. GEE models are appropriate for handling unbalanced longitudinal data (Ballinger, 2004). We present odds ratios (OR), 95% confidence intervals (CI), and *p* values. We assessed the unadjusted association between job insecurity and suicide risk and their association while controlling for potential confounders.

## Results

### Generalised estimating equation results

We ran independent GEE models with each of the proxy variables as a predictor and proxy suicide risk as the outcome. Each of the five proxy variables was significantly associated with higher suicide risk. As seen in Table 2, each proxy variable demonstrated a significant association with suicide risk. General health was the strongest predictor (OR = 5.10), followed by loneliness (OR = 4.81) and life satisfaction (OR = 4.35), supporting their inclusion in the proxy suicide risk score.

**Table 2.** Generalised Estimating Equation estimates for proxy predictors of suicide risk.

| Predictor | B | SE | OR | 95% CI | *p* |
| --- | --- | --- | --- | --- | --- |
| Subjective wellbeing – GHQ12 | 0.34 | .01 | 1.40 | 1.39, 1.41 | < .001 |
| Loneliness | 1.57 | .02 | 4.81 | 4.62, 5.00 | < .001 |
| General Health | 1.63 | .02 | 5.10 | 4.90, 5.31 | < .001 |
| Life Satisfaction | 1.47 | .02 | 4.35 | 4.18, 4.48 | < .001 |
| Current Financial Situation | 1.63 | .02 | 1.63 | 1.59, 1.67 | < .001 |

SE = Standard Error. OR = Odds Ratio. CI = Confidence Interval.

In the unadjusted GEE model, job insecurity was significantly associated with suicide risk. Compared to participants who were ‘very likely’ to lose their job (reference group), those who were ‘likely’ to lose their job had 36% lower odds of suicide risk (OR = 0.64, 95% CI: 0.54-0.75, *p* < .001), those who were ‘unlikely’ to lose it had 33% lower odds (OR = 0.67, 95% CI: 0.59-0.76, *p* < .001), and those who were ‘very unlikely’ to lose it had 22% lower odds (OR = 0.78, 95% CI: 0.69-0.89, *p* < .001).

In bivariate GEE models for suicide risk, in which we included job insecurity alongside individual social determinants, we found a change of 10% or greater in the odds ratio for job insecurity after adjusting for sex, ethnicity, marital status, education, and net personal income. These five variables were considered confounders in the relationship between job insecurity and suicide risk and were included in a final multivariable GEE.

In the multivariable GEE model, the association between job insecurity and suicide risk was not significant. Compared to the reference group, those who were ‘likely’ to lose their job had 5% lower odds of suicide risk (OR = 0.95, 95% CI: 0.81-1.10, *p* = .46), those who were ‘unlikely’ to lose it had 12% lower odds (OR = 0.88, 95% CI: 0.76-1.03, *p* = .10), and those who were ‘very unlikely’ to lose it had 9% lower odds (OR = 0.91, 95% CI: 0.76-1.09, *p* = .31).

In terms of covariates, we found significantly lower odds of suicide risk in males (6% lower odds; OR = 0.94, 95% CI: 0.89–0.99, *p* = .021), and participants with a degree (24% lower odds; OR⍰= ⍰0.76, 95% CI: 0.68–0.85, *p* < .001). Participants in the lowest income quartile had 68% higher odds of suicide risk (OR⍰= ⍰1.68, 95% CI: 1.50–1.79, *p*⍰< ⍰.001). Participants who were married or living as a couple had 40% lower odds compared to single participants (OR⍰= ⍰0.60, CI⍰= ⍰0.54–0.67, *p* < .001).

However, participants who were divorced (38%; OR⍰= ⍰1.38, CI⍰= ⍰1.20–1.54, *p* < .001) or separated (46%; OR⍰= ⍰1.46, CI⍰= ⍰1.25–1.70, *p* < .001) had higher odds. We found no significant differences for participants who were widowed (15% higher odds; OR⍰= ⍰1.15, CI⍰= ⍰0.90–1.35, *p* = .259). Although ethnicity was overall included in the model, none of the ethnic groups showed significantly higher association with suicide risk compared to the reference group (i.e., White British; all *p* > .05). Figure 1 summarises the odds ratios for the multivariable model.

**Figure 1.**
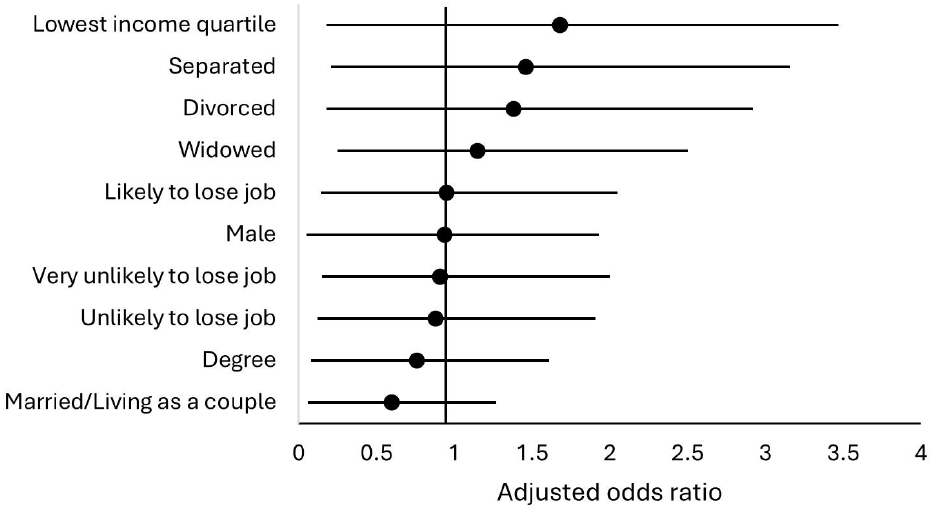
Adjusted odds ratios from multivariable Generalised Estimating Equation model **Figure 1 notes** Note. Adjusted for sex, ethnicity, marital status, education, and net personal income.

## Discussion

This study used longitudinal data from UKHLS to investigate the association between perceived job insecurity and suicide risk using a proxy score created from five psychosocial variables (i.e., subjective wellbeing, loneliness, general health, life satisfaction, and current financial situation). In line with past research (Blomqvist et al., 2024; Gallagher et al., 2025), the unadjusted and bivariate models, which included one social determinant at the time, showed a significant association between perceived job insecurity and suicide risk. However, when adjusting for sex, ethnicity, marital status, education, and net personal income, the association was no longer significant. Despite that, several covariates emerged as significant predictors. Being divorced or separated and being in the lowest income quartile were associated with higher odds of suicide risk. In contrast, being male, married or cohabiting, or holding a university degree were associated with lower odds of suicide risk. Overall, these findings highlight the multifaceted nature of suicide risk, which rarely stems from isolated factors but rather from the synergistic effects of multiple stressors (Franklin et al., 2017), and suggest that these variables may either protect against or exacerbate the psychological impact of job insecurity. While Durkheim (2005) emphasised macro-level social integration, the use of proxy suicide scores highlights the micro-level socioemotional disruptions that Abrutyn and Mueller (2014) argue are central to suicide risk. From a psychiatric perspective, this research highlights the importance of incorporating social determinants into suicide risk assessment, as traditional psychiatric evaluations may overlook the impact of financial difficulties, relationship instability, and educational disadvantage.

While job insecurity initially appeared to be a strong predictor of suicide risk, its effect is likely confounded by other social determinants. For example, having a university degree may protect from the impact of job insecurity by increasing employability, economic mobility, or equipping individuals with the necessary skills to cope or recover from job insecurity (Kim & von Dem Knesebeck, 2015). Being married or cohabiting may provide the required emotional support or reduce the impact of job insecurity due to shared financial resources (Næss, Mehlum, & Qin, 2021). Although males are more likely to commit suicide than females (WHO, 2025), females may be more vulnerable to the emotional effects of job insecurity (De Sio et al., 2018). These results suggest that these social determinants largely explain the relationship between job insecurity and suicide risk; therefore, job insecurity alone may not be a primary driver of suicide risk when broader contextual factors are taken into account.

The use of a proxy score, although necessitated by data limitations, was methodologically justified and validated by internal consistency and strong individual item associations. It was validated through an exploratory factor analysis by showing that all five variables loaded onto a single factor, supporting its use as a unidimensional measure of suicide risk.

Using a proxy allows for meaningful longitudinal exploration of suicide risk in datasets where direct measures are absent, and aligns with previous studies employing similar constructs (Borges et al., 2006; Carmel et al., 2016). It is a feasible method for identifying individuals at high psychiatric risk in large-scale datasets, and can inform population-based mental health surveillance and guide resource allocation to those with overlapping risk factors for suicide.

### Strengths and Limitations

Major strengths of the study included the use of a large nationally representative sample that allowed the investigation of variability over time. Applying Generalised Estimating Equations allowed for the analysis of unbalanced longitudinal data (datasets where participants contribute differing numbers of observations across waves), enhancing statistical power and generalisability of findings by enabling the inclusion of more cases. The study is not without limitations. Although the proxy score for suicide risk is supported by past research and was internally consistent, it is not a validated clinical tool and may not capture the complexity of suicide-related thoughts and behaviours, and may underrepresent clinical severity. In addition, removing a large portion of the sample due to missing data on job insecurity may have introduced bias, even though sensitivity analyses suggested otherwise.

### Implications and Future Research

The findings of this study suggest that policies targeting job insecurity alone may be insufficient to reduce suicide risk. Policies that promote mental health services or aim at reducing suicide risk should take into consideration broader social determinants such as financial inequalities, relationship status, and access to education. Interventions that protect from the effects of financial difficulties, support relationship stability, and provide educational opportunities for improving people’s skills are likely to be more effective than targeting perceptions of job insecurity. Integrating socioeconomic screening into primary care and psychiatric services could enhance early identification of high-risk individuals, while workplace mental health programmes could be adapted to address financial literacy, relationship stability, and coping strategies.

Future research should explore potential mediating mechanisms that may impact the relationship between job insecurity and suicide risk, such as chronic stress, social isolation, and accessing resources. Incorporating measures of suicidal ideation or attempts, coping strategies, and clinical markers could shed light on the causal pathways involved and expand this study’s findings. Future research should also explore how integrated screening models could be operationalised in clinical practice and assess interventions targeting social determinants. In addition, qualitative research may offer a better insight into individual experiences with job insecurity and the impact it has on their mental health.

### Conclusion

This study found that while perceived job insecurity initially appeared to predict suicide risk, its effect was explained by other social determinants, specifically income, marital status, and education. These findings highlight that suicide risk arises from a complex interplay of social, economic, and personal circumstances rather than single stressors. Addressing broader social determinants, such as reducing financial inequalities, strengthening social support systems, and promoting educational opportunities, may be more effective in mitigating suicide risk than targeting job insecurity alone. Future research should explore the mechanisms underlying these relationships to inform more targeted prevention strategies. Bridging epidemiological findings with primary care delivery will allow a more comprehensive, preventive approach to suicide that bridges public health and clinical psychiatry.

## Data Availability

All data produced are available online at UK Data Service

https://beta.ukdataservice.ac.uk/datacatalogue/studies/?Search=6614#!?Search=6614&Page=1&Rows=10&Sort=1&DateFrom=440&DateTo=2025

## Notes

### Competing Interest Statement

The authors have declared no competing interest.

### Funding Statement

This study did not receive any funding

### Author Declarations

We obtained the data from the UKHLS (www.understandingsociety.ac.uk; University of Essex, 2024). University of Essex (2024). Understanding Society: Waves 1-14, 2009-2023 and Harmonised BHPS: Waves 1-18, 1991-2009. [data collection]. 19th Edition. UK Data Service. SN: 6614, doi: 10.5255/UKDA-SN-6614-20

